# Interactions between seasonal temperature variation and temporal synchrony drive increased arbovirus co-infection incidence

**DOI:** 10.1101/2022.05.05.22274743

**Authors:** Marya L. Poterek, Chantal B.F. Vogels, Nathan D. Grubaugh, Gregory D. Ebel, T. Alex Perkins, Sean M. Cavany

**Affiliations:** Department of Biological Sciences, University of Notre Dame; Department of Epidemiology of Microbial Diseases, Yale School of Public Health, Yale University; Department of Microbiology, Immunology, and Pathology, Colorado State University

## Abstract

Though instances of arthropod-borne (arbo)virus co-infection have been documented clinically, the overall incidence of arbovirus co-infection and its drivers are not well understood. Now that dengue, Zika, and chikungunya viruses are all in circulation across tropical and subtropical regions of the Americas, it is important to understand the environmental and biological conditions that make co-infections more likely to occur. To understand this, we developed a mathematical model of cocirculation of two arboviruses, with transmission parameters approximating dengue, Zika, and/or chikungunya viruses and co-infection possible in both humans and mosquitoes. We examined the influence of seasonal timing of arbovirus cocirculation on the extent of co-infection. By undertaking a sensitivity analysis of this model, we examined how biological factors interact with seasonality to determine arbovirus co-infection transmission and prevalence. We found that temporal synchrony of the co-infecting viruses and average temperature were the most influential drivers of co-infection incidence. For seasonal patterns typical of a tropical region, we observed non-negligible incidence irrespective of arrival time when two arboviruses arrived simultaneously. Under our default parameter settings, this corresponded to a maximum co-infection cumulative incidence of 83 per 1,000 individuals and a minimum cumulative incidence of 32 per 1,000 individuals in the year following arrival. For seasonal patterns typical of a more temperate region, co-infections only occurred if arrivals took place near the seasonal peak, and even then, did not reach 0.01 co-infections per 1,000 individuals. Our model highlights the synergistic effect of co-transmission from mosquitoes, which leads to more than double the number of co-infections than would be expected in a scenario without co-transmission. Our results show that arbovirus co-infections are unlikely to occur in appreciable numbers unless epidemics overlap in space and time and in a tropical region.

## INTRODUCTION

The past decade has seen the Americas affected by epidemics of both Zika and chikungunya, adding to the burden of arthropod-borne (arbo)viral disease in a region where seasonal dengue epidemics were already a regular occurrence in most countries (1–3). All three of the viruses that cause these diseases are spread by the same vectors: *Aedes aegypti* and *Aedes albopictus* mosquitoes. Hence, the diseases’ spatiotemporal distribution is largely determined by the same environmental and climatological drivers (4–7). This has led to overlapping epidemics of two and three viruses, which in turn has led to many reports of co-infections (8). The rate of co-infections with multiple arboviruses is magnified by the ability of the vector to be simultaneously co-infected with two or more viruses and to co-transmit two or more viruses with a single bite (9).

The phenomenon of arbovirus co-infection is still largely understudied with many unknowns (8). For instance, while some studies have reported an increased risk of severe outcomes in co-infections of dengue virus (DENV) and chikungunya virus (CHIKV), other studies have not observed this (10,11). Similarly, while co-infection involving Zika virus (ZIKV) does not alter the clinical presentation of uncomplicated infections, it is unclear whether it alters the risk of severe disease (12). It is also unclear the extent to which prior or recent infection with one virus can enhance or protect against subsequent infection with another (13–16). When multiple arboviruses circulate in the same region at the same time, the combination of uncertainty about cross-protection versus mutual enhancement, differing importation times of each virus, and strong seasonal climate drivers, leads to potentially complex temporal patterns of single infection and co-infection (17).

Seasonal climate drivers play an important role in arbovirus infection dynamics, as variations in temperature determine environmental suitability for mosquito vector survival and virus transmission (18–21). Arbovirus epidemic size and duration are a product of both mean temperatures and seasonal variation and are maximized under conditions that promote mosquito survival (22). Tropical climates are generally more suitable for arbovirus vectors (6) and are therefore more likely to experience recurring arbovirus epidemics, which leads to the accrual of immunity in human populations who live there (23). While much remains unknown about the level of cross-immunity between arboviruses, preexisting immunity in a population is likely to impact the dynamics and observed patterns of arbovirus co-infections, as well.

Data on the frequency of arbovirus co-infection remain sparse (8) and where data does exist there are many factors which could lead to variability between studies, such as cross-immunity, epidemic timing, and seasonality. In this context, mathematical modeling provides a useful way to synthesize our understanding of arbovirus transmission and explore the conditions which may most likely give rise to a heightened burden of arbovirus co-infection. To do this, we built a temperature-dependent mathematical model of arbovirus co-circulation and co-transmission that permits cross-protection between arboviruses and asynchronous epidemics. We first use the model to understand the interplay of differing importation times and seasonal transmission in an immunologically naive population. Next, we describe how the burden of co-infection could change under differing levels of immunity and cross-protection. Finally, we undertake a global sensitivity analysis of our model’s parameters to provide a holistic view of the conditions which may lead to the highest frequency of co-infection in humans.

## METHODS

### Model

We used a deterministic SEIR-SEI model to explore the influence of temperature on arbovirus co-infection magnitude and timing. This model incorporates two arboviruses, referred to as virus A and virus B, with identical transmission and human recovery rate parameters. We relied upon several of the structural assumptions and parameter values reported by Vogels et al. (8), particularly those governing co-transmission. In our model, transmission from co-infected humans and mosquitoes occurs with the same probability as transmission from singly-infected humans and mosquitoes, and there is no mortality effect from infection in either humans or mosquitoes (9,24). Consistent with Rückert et al. (9), we assume that 60% of mosquitoes become co-infected following a blood meal on a co-infected human, while 20% become singly-infected with virus A and 20% singly-infected with virus B. Our model implements an intermediate transmission scenario, in which 50% of bites from a co-infected mosquito lead to co-infection and 50% lead to a single infection, the latter split evenly between the two arboviruses. We assessed model sensitivity to this assumption of intermediate transmission, which has been used in previous modeling work (8). Viruses A and B have identical, dengue-like parameters (Table 2).

We additionally made several assumptions about the roles that exposed and infected individuals play in transmission. Co-infection in humans and mosquitoes can occur via co-transmission to susceptible individuals or sequential transmission involving individuals who are susceptible and then exposed to the first of two viruses. Following their infectious period, individuals recover, and if they have only been singly infected by one virus can then be singly infected with the other virus. Infection while an individual is exposed restarts the incubation period. In our baseline analysis, we assumed no cross-protection or enhancement from a prior infection but explored this in later analyses. Structural details of the model are illustrated in Figure 1, with sequential transmission indicated in red and co-transmission in blue.

**Figure 1:**
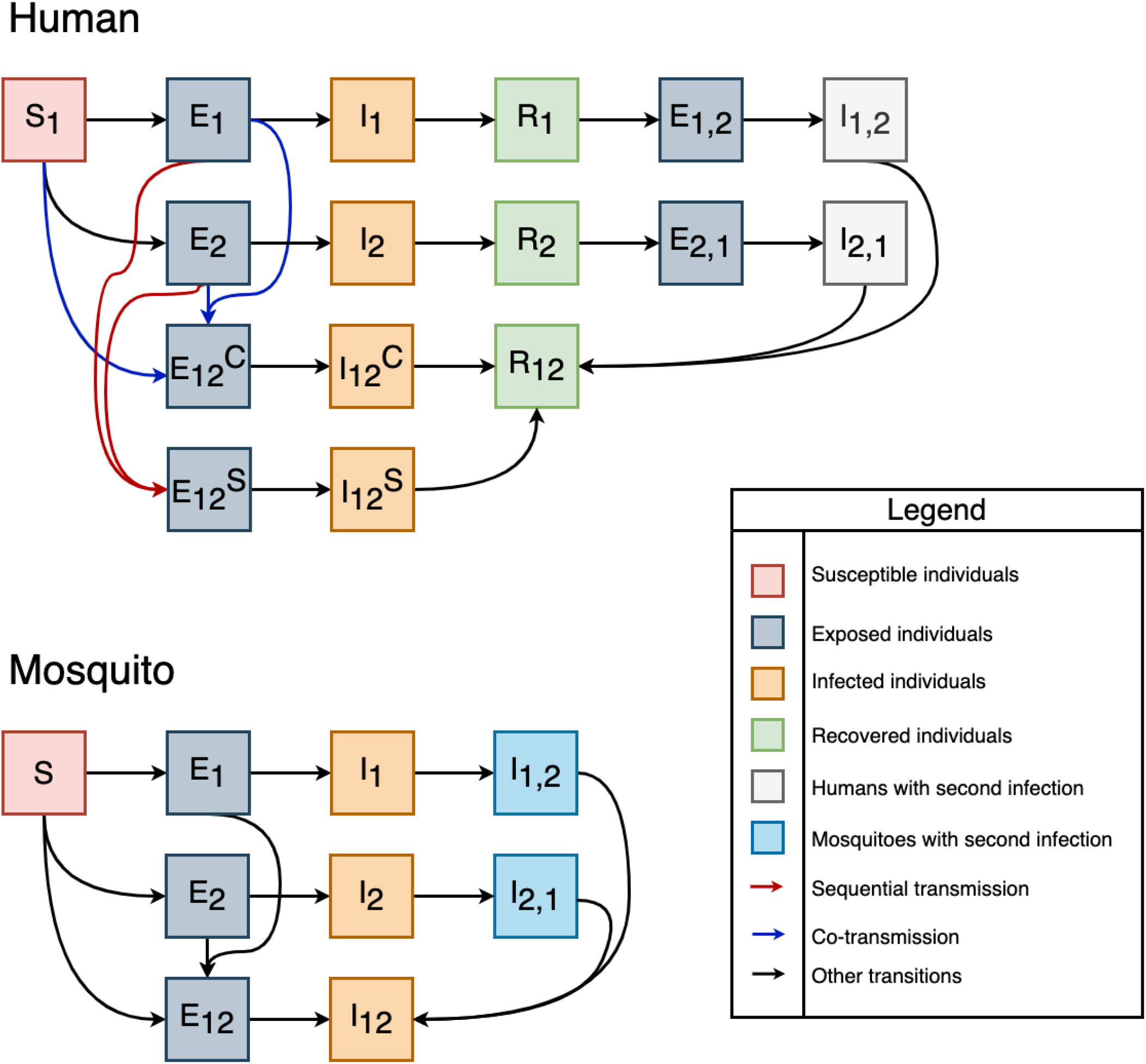
Model diagrams for the human and mosquito components of the model. Red arrows indicate sequential transmissions into a co-infected state, while blue arrows indicate co-transmissions into a co-infected state. Superscripts refer to co-transmission (C) or sequential transmission (S); co-transmission can occur to susceptible or exposed individuals. Subscript “12” refers to co-infection, and “1,2” refers to secondary infection with the second virus following recovery from the first. Compartments are colored by state.

We incorporated a seasonal component into the model by using temperature-dependent parameters, where biologically appropriate (Table 1). We used sinusoidal temperature curves with a period of one year to drive the values of these parameters, with mean and amplitude chosen to reflect specific climate regimes (Rio de Janeiro for tropical regions, Beijing for temperate regions). We did not consider diurnal temperature variation. Temperature-dependent parameters were those describing *Aedes aegypti* life history: mosquito biting rate, probability that an infected mosquito transmits to a human during feeding, probability that a mosquito becomes infected after feeding on an infected human, mosquito mortality rate, and virus extrinsic incubation rate. Their values were chosen with reference to previous modeling work on fitted thermal responses for *Aedes aegypti* (22). This approach allowed us to explore the relative influences of seasonal timing and temporal synchrony on arbovirus co-infection under several climate scenarios.

**Table 1:** Temperature-dependent parameters describing fitted *Aedes aegypti* life traits and arbovirus transmission (22).

| Parameter | Definition | Function | Fitted Parameters |  |  |
| --- | --- | --- | --- | --- | --- |
| a | Mosquito biting rate | Briere | c=13.4 | $T_{\min} = 40.1$ | $T_{\max} = 2.0 \times 10^{-4}$ |
| b | Probability that an infected mosquito transmits to a human during feeding | Briere | c=17.1 | $T_{\min} = 35.8$ | $T_{\max} = 8.5 \times 10^{-4}$ |
| c | Probability that a mosquito becomes infected after feeding on an infected human | Briere | c = 12.2 | $T_{\min} = 37.5$ | $T_{\max} = 4.9 \times 10^{-4}$ |
| g | Mosquito mortality rate | Quadratic | c = 9.2 | $T_{\min} = 37.7$ | $T_{\max} = -1.5 \times 10^{-1}$ |
| $\epsilon^M$ | Virus extrinsic incubation rate | Briere | c = 10.7 | $T_{\min} = 45.9$ | $T_{\max} = 6.7 \times 10^{-5}$ |

**Table 2:** Temperature-independent parameters.

| Parameter | Definition | Value | One-at-a-time sensitivity analysis range | Source(s) |
| --- | --- | --- | --- | --- |
| $\alpha$ | Coefficient of cross-protection | 0-1, varied | 0-1 | n/a |
| $r$ | Average time for a human to recover | 5 days | 3-10 days | (30–32) |
| $p_{12}$ | Probability that when a co-infected mosquito transmits, it transmits both viruses to a human | 0.5 | 0-1 | (8) |
| $p_1$ | Probability that when a co-infected mosquito transmits, it transmits only virus X to a human | 0.25 | $0.5(1 - p_{12})$ | (8) |
| $p_2$ | Probability that when a co- infected mosquito transmits, it transmits only virus Y to a human | 0.25 | $0.5(1 - p_{12})$ | (8) |
| $p_1^M$ | Probability that when a mosquito becomes infected after feeding on a co-infected human, it only becomes infected by virus X | 0.2 | n/a | (8,9) |
| $p_2^M$ | Probability that when a mosquito becomes infected after feeding on a co-infected human, it only becomes infected by virus Y | 0.2 | n/a | (8,9) |
| $p_{12}^M$ | Probability that when a mosquito becomes infected after feeding on a co-infected human, it becomes infected by both viruses | 0.6 | n/a | (8,9) |
| $\epsilon^H$ | Incubation period (human) | 7 days | 4-8 days | (28,29,33) |

#### Equations

Model equations for humans are as follows, with parameter values and meanings shown in Tables 1-2. Eqs. 1-3 define the forces of infection for each infection for each virus and for co-transmitted viruses, which involves input from mosquito states and transmission probabilities. Eqs. 7-8 and 11-12 distinguish between co-transmitted co-infections, in which individuals become infected with two arboviruses simultaneously, and sequentially-transmitted co-infections, in which individuals initially infected with a single arbovirus become infected with a second. Eqs. 16-19 describe the process of acquiring a second infection for individuals who have completely recovered from an initial infection, with possible cross-protective immunity.

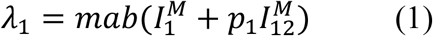

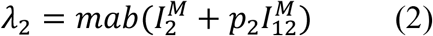

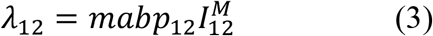

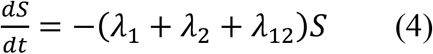

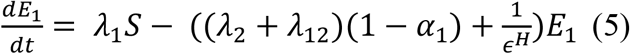

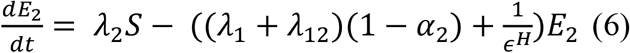

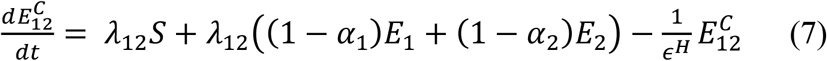

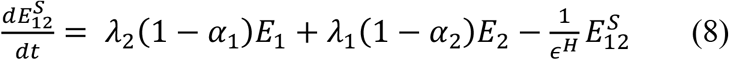

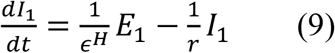

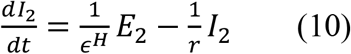

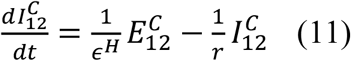

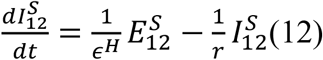

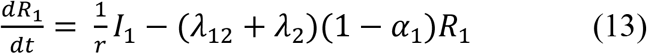

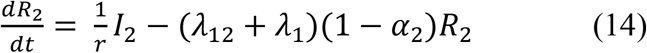

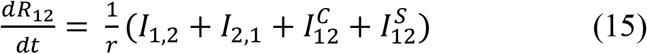

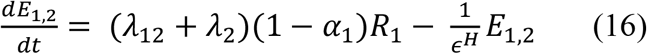

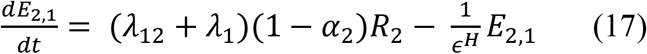

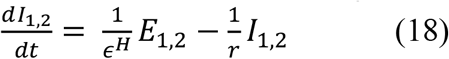

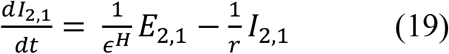

Model equations for mosquitoes are as follows, with parameter values and meanings shown in Tables 1-2. Eqs. 20-22 define the forces of infection for each infection for each virus and for co-transmitted viruses, which involves input from human states and transmission probabilities. Eqs. 26 and 29 describe mosquito co-infection, which is driven by temperature-dependent parameters. Eqs. 30-31 outline how mosquitoes in this model become infected with a second arbovirus after recovering from their first infection.

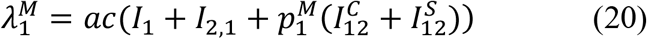

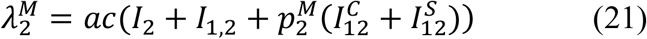

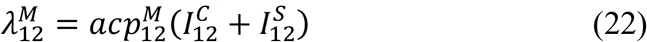

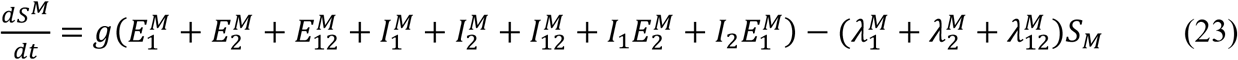

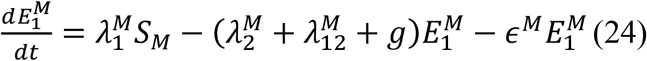

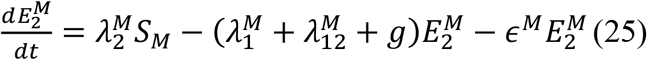

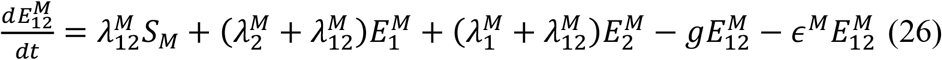

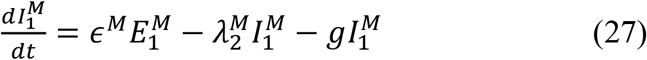

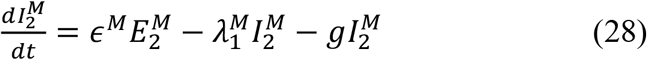

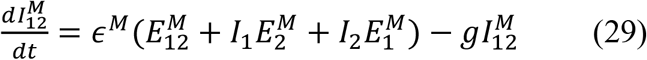

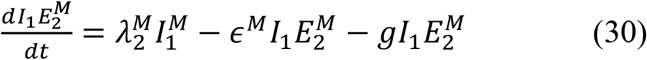

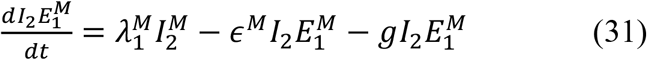

Equations to address seasonal fluctuations in temperature and thermal traits across a 365-day period are as follows, modeled after the approach to seasonal forcing used by Huber et al. (22). In Eq. 32, *T*_*max*_, *T*_*min*_, and *T*_*mean*_ represent the maximum, minimum, and mean temperature for a region across a calendar year. In Eqs. 33 and 34, *c, T*_*max*_, *T*_*min*_, and *T* represent the fitted rate constant, critical temperature maximum, critical temperature minimum, and temperature at a given time, respectively. As in Mordecai et al., we assumed that values above the critical maximum and below the critical minimum were zero (25).

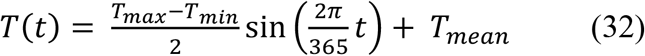

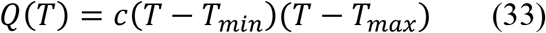

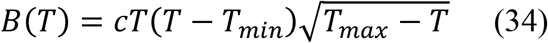

#### Parameters

Fitted parameters describing *Aedes aegypti* life traits and arbovirus transmission are shown in Table 1. Temperature dependence of traits was described using quadratic or Briére functions and fitted to experimental data (22,25). The value of the parameters describing these traits varies in our model as seasonal temperatures fluctuate.

Additional population-level parameters were governed by temperature. To ensure that the ratio of mosquitoes to humans, *m(T)*, remained biologically feasible regardless of climate, we followed the approach of Siraj et al. (26) and developed a mosquito ratio scaling factor, *γ*, such that

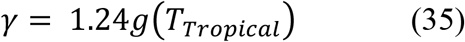

and

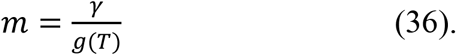

*γ* is defined here as the product of the estimated ratio of mosquitos to humans in Rio de Janeiro in 2012 (1.24) and the temperature-varying fitted mosquito mortality rate at temperatures typical of that city, such that *T*_*Tropical*_ = 24.3 (Eq. 35) (22,26,27). Using this value to scale *m* across various temperature environments ensured that the ratio of mosquitoes to humans remained biologically feasible, between 1.18 and 1.25 over the course of each simulation (Eq. 36).

Temperature-independent parameter values and definitions are consistent with those in Vogels et al. (8), which developed a generic model of arbovirus co-infection that ours is built on. Transmission parameters pertaining to *Aedes aegypti* mosquitoes follow those used in previous dengue modeling studies (16,28,29) while transmission parameters pertaining to co-transmission from co-infected humans to mosquitoes were informed by data from Rückert et al. (9).

### Analyses

#### Outcomes of interest

We focused on four model outputs: 1) cumulative incidence of infection with virus A, 2) cumulative incidence of infection with virus B, 3) cumulative incidence of co-infection, and 4) proportion of all infections that were co-infections. All quantities were defined as cumulative values across the course of a year-long simulation, at which time all arbovirus outbreaks had run their course.

#### One-at-a-time sensitivity analysis of temperature-independent parameters

While the majority of the parameters governing arbovirus infection and co-infection in our model were temperature-dependent, three were not: cross-protection (α), recovery time (*r*), and human incubation period (ε^H^). We considered the individual impact of these parameters on model outputs in a series of one-at-a-time sensitivity analyses. For each parameter, we varied the value across a plausible range (Table 2) while holding the remaining two temperature-independent variables constant and examined the relationship between the varied parameter and selected model outputs. We then repeated this analysis under several assumptions about population-level preexisting immunity—25% immunity to virus A, 25% immunity to virus B, and 25% immunity to both—and considered the aforementioned model outputs’ response to these population scenarios.

#### Explore differing roles of importation time and seasonality

The seasonal component of the model made it possible to examine the effect of seasonal temperature variation on the cumulative incidence of co-infection, as well as the effect of temporal synchrony or asynchrony of the co-infecting viruses. We evaluated this effect under two temperature regimes, defined by mean temperatures and seasonal amplitudes for a given region and based upon 2019 monthly mean values obtained from Weather Underground (wunderground.com). These included an environment with temperatures typical of a tropical region (mean 25.1 °C, amplitude 3.4 °C; similar to Rio de Janeiro), and an environment with temperatures typical of a more temperate region (mean 13.8 °C, amplitude 14.7 °C; similar to Beijing). Using monthly mean temperatures for the two cities, we calculated associated mean, minimum, and maximum temperatures across a year to determine the average temperature and seasonality observed in the most recent full calendar year. We systematically considered each possible combination of virus arrival times within a simulation and compared simulation results between the two temperature settings.

#### Global sensitivity analysis

To evaluate the interaction components of our model parameters, we conducted a global, variance-based sensitivity analysis, also known as a Sobol sensitivity analysis, using the SALib library in Python (34). This approach is not dependent on the presence of monotonic relationships between input parameters and outputs. With this analysis, we were able to quantify the amount of variance in the aforementioned model outputs that could be attributed to individual input parameters, as well as the amount of variance that could be attributed to pairwise and higher interactions among these parameters. We varied all temperature-independent parameters and the mean and amplitude of the yearly temperature curve in this analysis. We used the Saltelli sampling scheme to generate 1.8 million parameter combinations from a range of plausible values (Table 3) to ensure that we covered the parameter space of biological interest. Our sensitivity analysis was conducted on the corresponding 1.8 million model outputs, once for each of four population immunity scenarios: no existing immunity, 25% existing immunity to virus A, 25% existing immunity to virus B, and 25% existing immunity to both infections.

**Table 3:** Sampling ranges for global sensitivity analysis.

| Parameter | Sampling Range |
| --- | --- |
| Mean temperature | 0 – 36 °C |
| Mean amplitude | 0 – 20 °C |
| Date of importation of virus A | 0 – 365 |
| Human recovery time ($r$) | 3 – 10 days |
| Human incubation period ($\epsilon^H$) | 5 – 8 days |
| Virus introduction interval | 0 – 75 days |
| Coefficient of cross-protection ($\alpha$) | 0 – 1 |
| Probability of human co-transmission ($p_{12}$) | 0 – 1 |

## RESULTS

### The role of virus importation timing, seasonality, and temperature

To better understand what combinations of epidemic timing and seasonality lead to a high incidence of co-infections, we explored a range of virus arrival times throughout the year under tropical and temperate temperature regimes. This scenario is reflective of a situation where two arboviruses are imported to a location within a year of each other, as happened with Zika and chikungunya viruses in some South American countries in the 2014-2016 period. When seasonal patterns resembled those in a tropical region (25.1 °C, amplitude 3.4 °C), simultaneous (same-day) virus importation resulted in incidence of co-infection that was always greater than 19 per 1,000 individuals, although seasonal differences were observed (Fig. 2A-B). Simultaneous arrival of viruses A and B resulted in co-infection incidence ranging from 32 to 83 per 1,000 individuals per year, with low incidence being associated with periods of significant negative temperature change, particularly in late summer (days 100-150) (Fig. 2A). Asynchronous virus arrival resulted in fewer co-infections than simultaneous arrival did, with larger gaps between virus arrival dates corresponding to lower incidence of co-infection (Fig. 2B).

**Figure 2:**
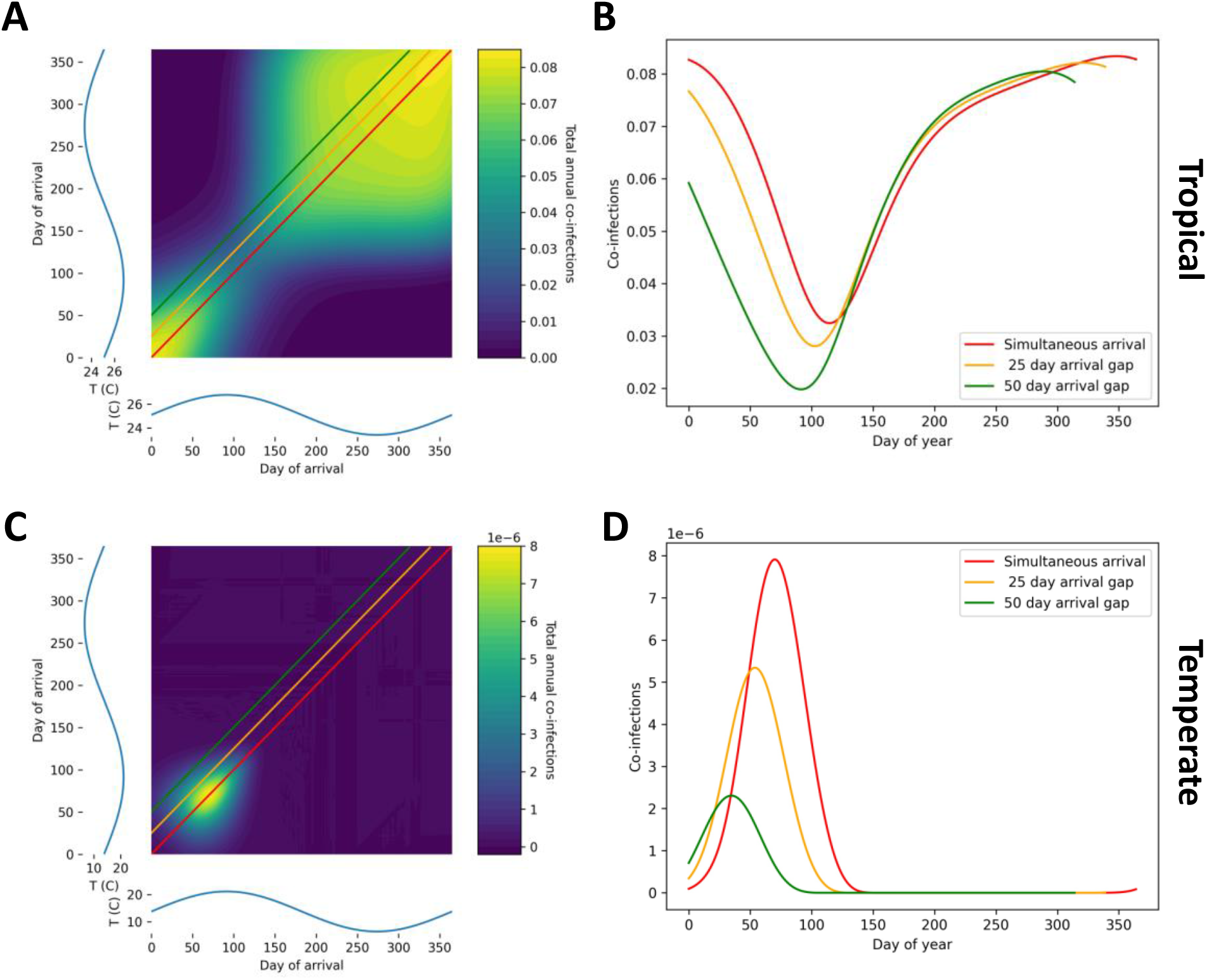
Seasonal temperature and co-infection responses under different temperature regimes. Fill colors indicated total co-infections observed in a year (or the attack rate), rather than an instantaneous measurement, given each possible combination of virus importation dates. Temperature curves are shown in blue on each axis of the contour plots, and approximate those in Rio de Janeiro (A-B, 25.1°C, amplitude 3.4°C), and those in Beijing (C-D, mean 13.8°C, amplitude 14.7°C). Colored lines on each plot show how the attack rate observed in each year changes given different intervals between virus importation dates.

In contrast, when seasonal patterns resembled those of a more temperate region (mean 13.8 °C, amplitude 14.7 °C), both simultaneous virus arrival and seasonal high temperatures were required to observe non-negligible co-infection, with a maximum incidence of 0.0079 per 1,000 individuals (Fig. 2C-D). Asynchronous arbovirus arrival resulted in seasonal trends in co-infection consistent with simultaneous arrival but produced negligible co-infections as the time between arrivals grew (Fig. 2D). Arrivals after the summer (around day 100) generated similarly negligible co-infection incidence, as temperatures fell below those conducive to virus transmission by *Aedes aegypti* mosquitoes (Fig. 2C).

In addition to being temperature-driven, model outputs are influenced by several biologically important temperature-independent parameters, including immunological cross-protection, recovery time, and the human incubation period. The coefficient of cross-protection (α) refers to the level of protection each infection provides against the other, where 0 indicates no protection, and 1 indicates complete protection. When values of α increased, the incidence of virus A decreased slightly, and the proportion of co-infections decreased dramatically (Figs. 3A, D). This decrease in the incidence of virus A when cross-protection was high was driven by individuals with virus B who experienced a reduced force of infection of virus A and was limited by the later arrival of virus B into the population. The proportion of co-infections declined steeply as high values of α inhibit both sequential and co-transmitted co-infections from occurring. However, high α favors co-transmitted co-infections over sequentially transmitted co-infections because co-transmission is limited by cross-protection to the extent that sequential transmission is.

**Figure 3:**
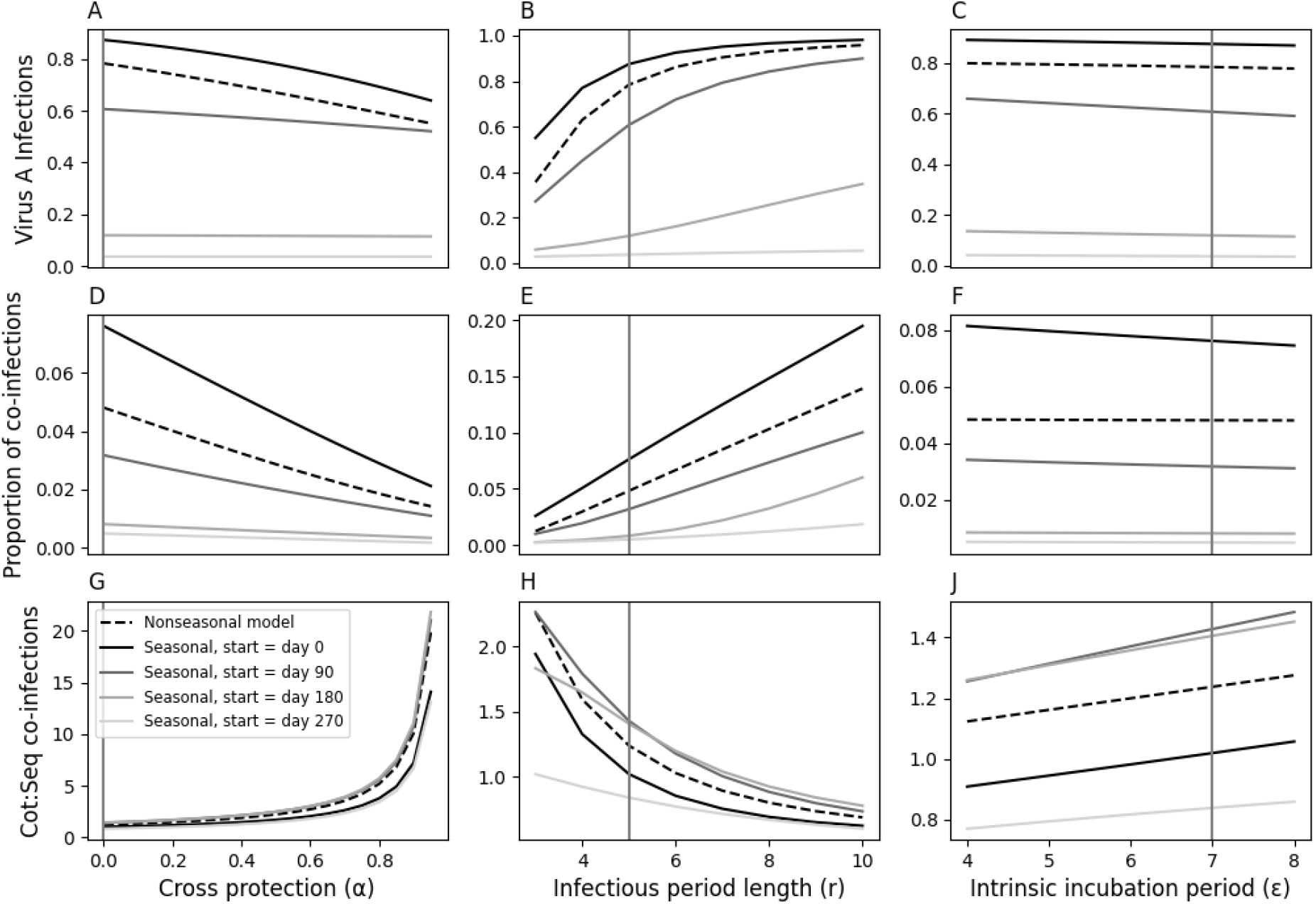
Univariate sensitivity to temperature-independent parameters under seasonal and non-seasonal models. Varying initial importation dates of virus A were considered when the seasonal model was used to explore the range of temperature environments possible within a year. Vertical lines indicate the baseline value for each parameter, and y-axes differ for each subplot. Panels D-F show the proportion of all infections that are co-infections. Panels G-J show the ratio of co-transmitted co-infections to sequentially-transmitted co-infections.

Model outputs are also noticeably influenced by changes in the value of recovery time (*r*), the average time in days it takes for a human to recover from either infection. Longer recovery times correspond to higher virus A incidence and a higher proportion of co-infections, as more time spent infectious allows for greater exposure to a second infection and increases the reproduction number of both viruses (Figs. 3B, E). Longer recovery times lead to a lower ratio of co-transmitted co-infections to sequentially transmitted co-infections for the same reason—time spent infectious, where another infection cannot be acquired immediately, favors sequential infection transmission (Fig. 3H).

In contrast with the previous two parameters, model outputs do not appear particularly susceptible to changing incubation period values. Human incubation period (ε^H^) is also measured in days, and has been approximated in studies of dengue and Zika virus to be 5-8 days (28,35). Within this range, ε^H^ does not strongly influence the incidence of either single or co-infections (Figs. 3C, F). However, the seasonal and non-seasonal models produce noticeably different model outputs observe noticeable differences in seasonal model outputs depending on the day virus A is imported (Figs. 3C, F, I). When virus A was imported in fall or winter, the incidence of virus A and the proportion of infections that were co-infections were relatively low as compared to spring or summer importation days. While both the seasonal and non-seasonal models have parameters that approximate values for a tropical-like region, earlier importation dates within the year appear more suitable for arbovirus infection and co-infection, as rising temperatures following importation reach values ideal for transmission once incidence has grown.

### Role of preexisting immunity

In many tropical environments, transmission of some arboviruses occurs in semi-regular seasonal cycles, which will impact the incidence of co-infection as part of the population will have prior immunity to one or more of the viruses. To consider how this might influence co-infections, we examined the behavior of the model with respect to four scenarios about initial conditions for population immunity: 1) none; 2) 50% immune to virus A; 3) 50% immune to virus B; and 4) 50% immune to both viruses (the same 50% of the population immune to virus A was also immune to virus B) (Fig. 4). Scenario 2, in which virus B is introduced to a population with some immunity to virus A, reflects patterns in immunity similar to those observed when Zika or chikungunya viruses have been introduced in dengue-endemic settings. We used the non-seasonal model for this analysis, and we set the time between importation of the two viruses to 30 days. Results from the baseline scenario were equivalent to those from the non-seasonal scenario in Figure 3.

**Figure 4:**
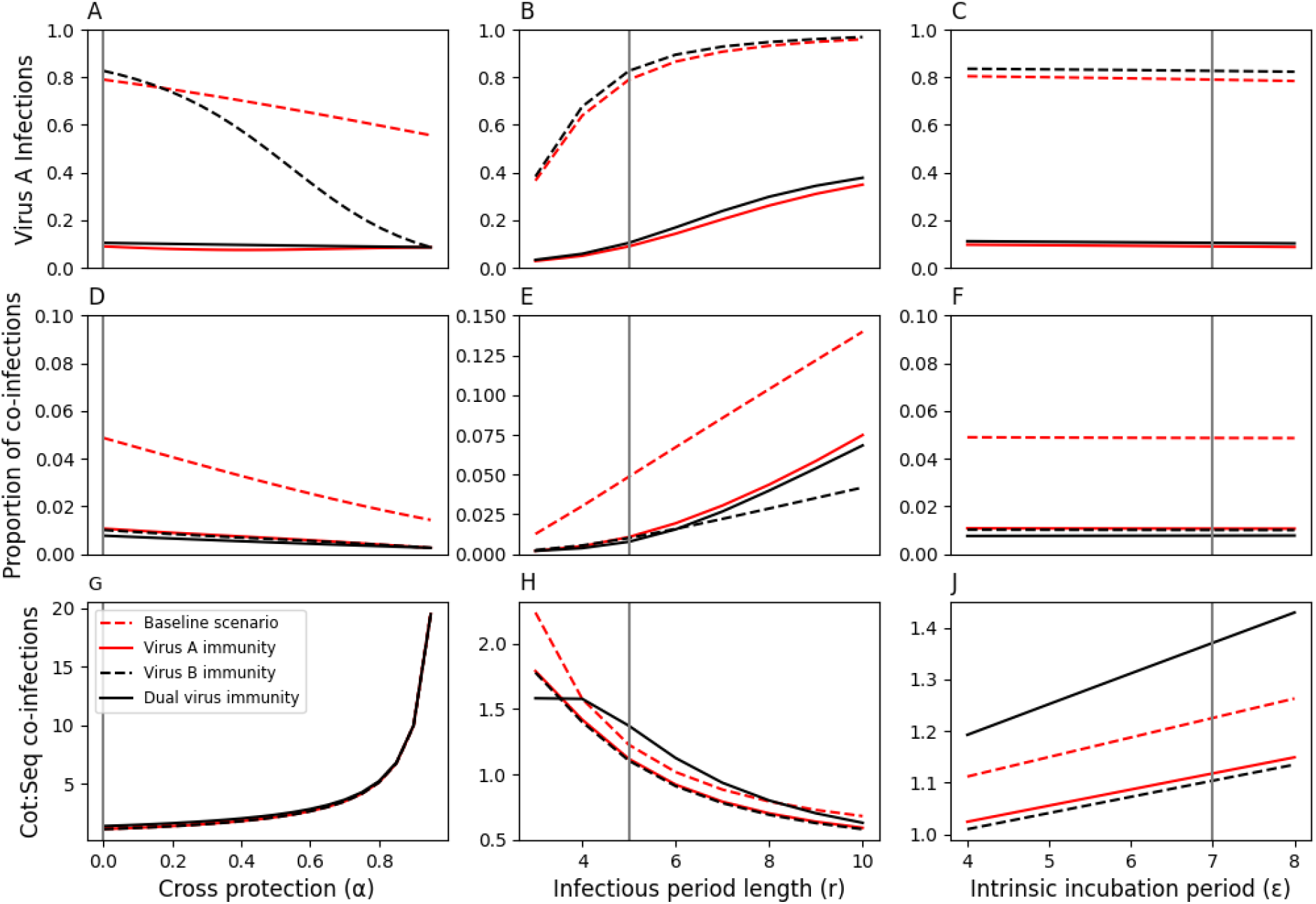
Non-seasonal model output in response to varied temperature-independent parameters under different initial immunity conditions. In each immunity scenario, 50% of the population are immune to a given virus or viruses, while the baseline immunity scenario includes no preexisting immunity. Vertical lines indicate the baseline value for each parameter. Panels D-F show the proportion of all infections that are co-infections. Panels G-J show the ratio of co-transmitted co-infections to sequentially-transmitted co-infections.

When high levels of cross-protection were present, immunity to virus B limited infection by either virus once virus B became prevalent (Figs. 4A, D). This resulted in a decrease in virus A incidence driven by the lower proportion of individuals not immune to virus B and, thereby, partially immune to virus A. In contrast, scenarios with immunity to virus A and immunity to both produced negligible incidence of virus A infections (Figs. A, D), since both limited the population susceptible to virus A.

Preexisting immunity had the most noticeable impact on the ratio of co-transmitted co-infections to sequentially transmitted co-infections, where we observed that co-transmitted co-infections were more heavily represented under the scenario with immunity to both viruses (black line) than they were under the no-immunity scenario (red dashed line) (Fig. 4J). When there was no initial immunity, there was a larger group of individuals susceptible to virus A at the beginning, which provided more opportunities for sequentially-transmitted co-infections. Since that population of individuals was much smaller when there was immunity to both viruses at the beginning, sequential transmission occurred less frequently. More generally, though, immunity of any type led to a much smaller proportion of infections that were co-infections (Figs. 4D-F). While this proportion was low even under the no-immunity scenario, immunity to even a single virus limited the occurrence of both sequential and co-transmitted co-infections.

### Variance-Based Sensitivity Analysis

To gain a holistic view of the effect of the examined biological and environmental factors and their interactions on the epidemiology of arbovirus co-infection, we used a variance-based sensitivity analysis. This approach allowed us to explore the contribution of each parameter to the variance of each model output. Examining the first-order indices, or those describing the direct relationship between each parameter and each model output, revealed that interactions between parameters accounted for more than 50% of the total variance in all four outputs (Fig. 5). However, when total-order indices were considered, which measure all contributions of input parameters to output variance (including interactions), parameters related to temperature and timing had the greatest effect on the outputs, especially temperature amplitude, temperature mean, the importation date of virus A, and the importation date of virus B. Together, these four parameters accounted for 85% of the total variance in the proportion of co-infection we observed, implying that the timing of outbreaks is by far the most important determinant of the level of co-infection. Additionally, the parameter governing the time between arbovirus introductions (virus B importation date) was influential on co-infection-related model outputs when there was preexisting immunity to virus B, as larger intervals between virus importation times could severely limit the potential for any temporal overlap between the viruses (Supp. Figs. 2-3). We also observed that the coefficient of cross-protection had a much smaller effect on incidence of virus A than it did on the other three outputs, due to interactions with the parameters responsible for temperature and its timing within the year (Fig. 5, Supp. Figs. 1-3).

**Figure 5:**
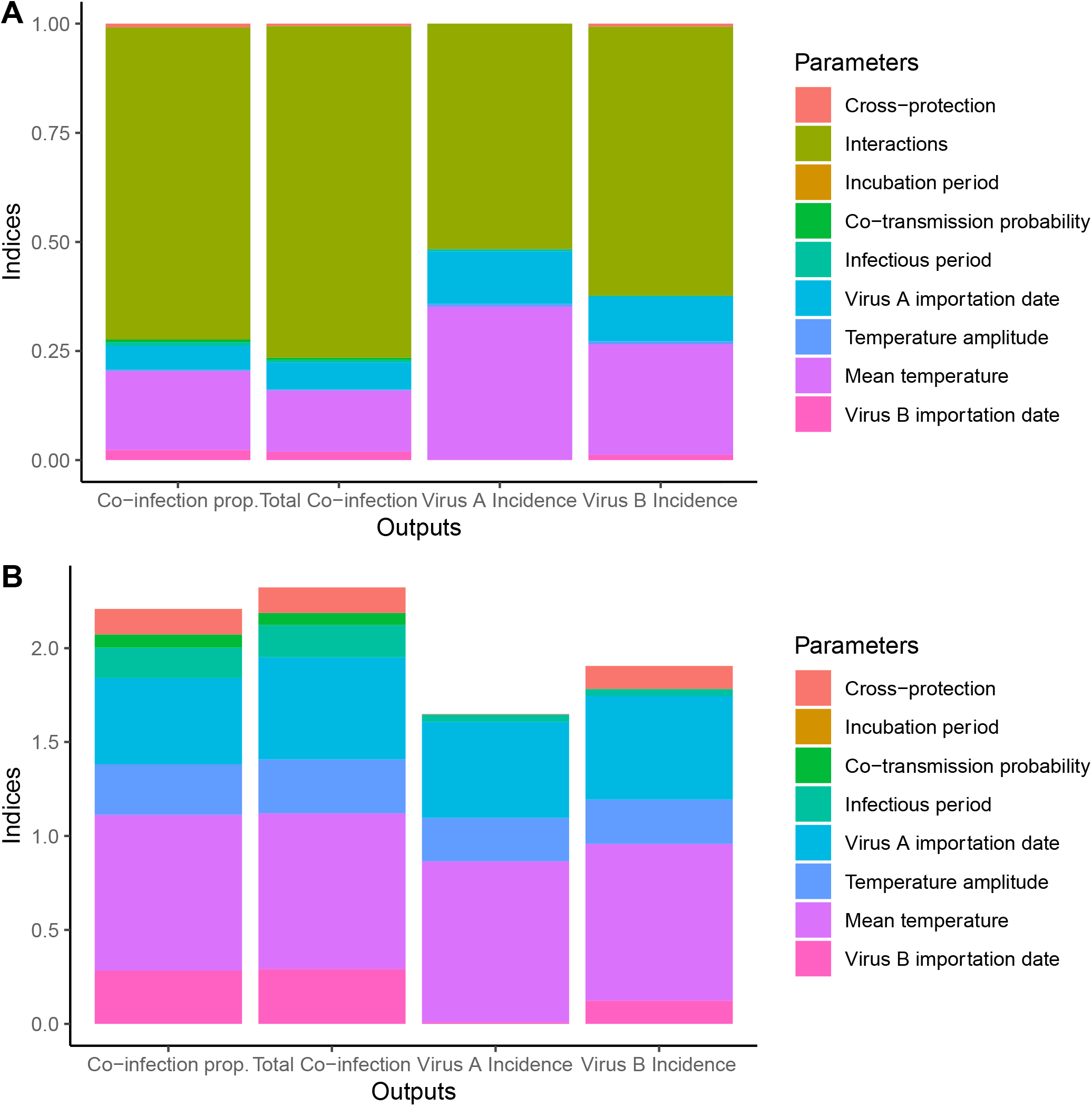
Output of variance-based sensitivity analysis for baseline immunity scenario (no preexisting immunity). Both first-order (A) and total-order (B) indices are shown. Fill colors indicate the parameter responsible for a given fraction of the variance in a given output. First-order indices sum to one, while total-order indices additionally account for all variance caused by a parameter’s interactions and therefore have no such constraint.

We further explored the interactions between parameters that contribute to variance in the incidence of co-infection and found these relationships to be consistent with the results from first- and total-order indices alone (Fig. 6). Interactions between mean temperature and other timing and temperature-related parameters, particularly virus A importation date, explained much more variance in this output than did other parameter combinations.

**Figure 6:**
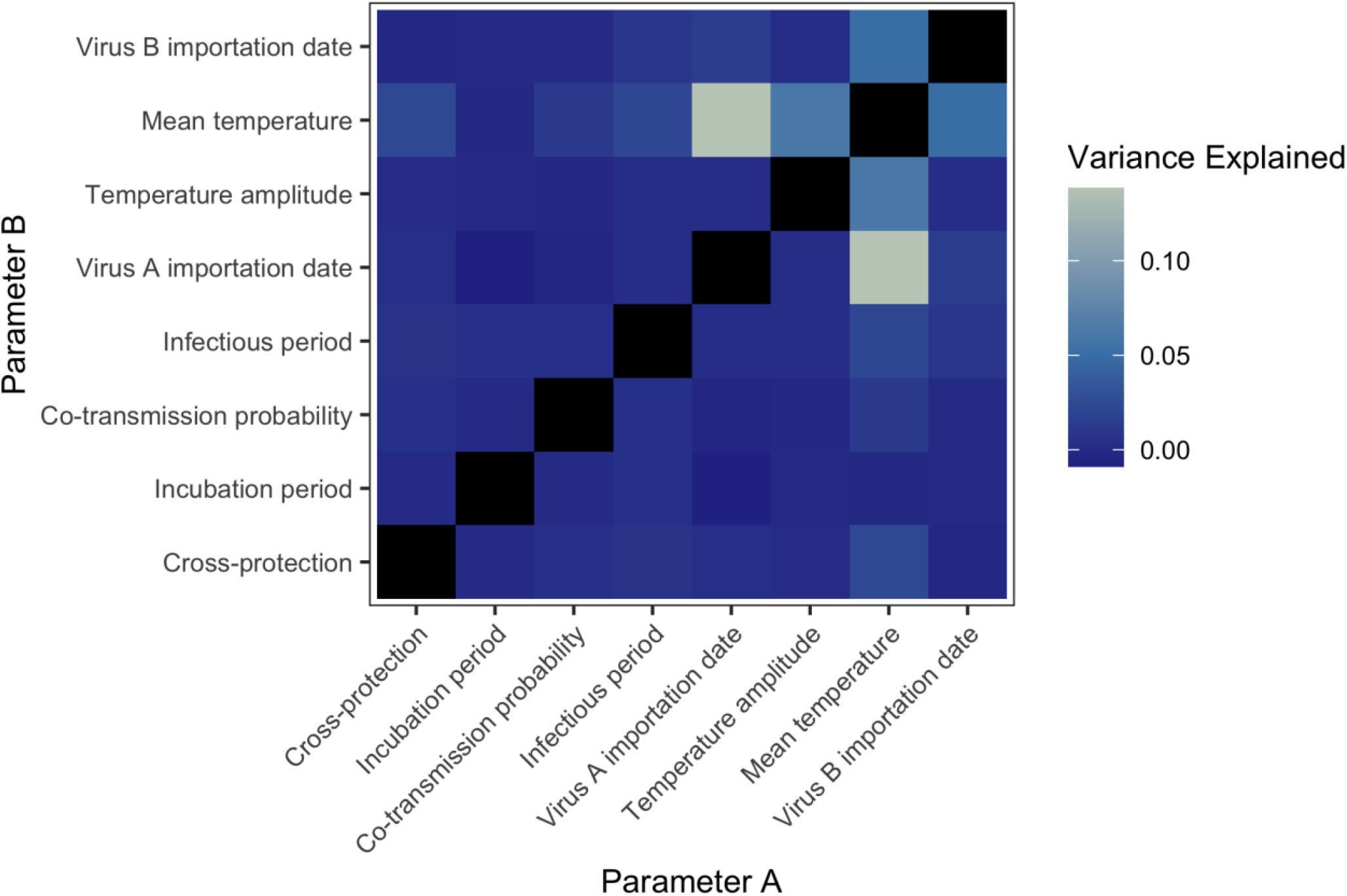
Total-order variance-based sensitivity analysis parameter interactions for total co-infection output under the baseline immunity scenario (no preexisting immunity). Black boxes on the diagonal represent first-order interactions, which were not considered here.

Additionally, we assessed the role that interactions between the viruses themselves play in observed co-infections, particularly via the mechanism of co-transmission. As our model allows for simultaneous arbovirus co-transmission between mosquitoes and humans, a phenomenon documented frequently in laboratory studies (9), the frequency of arbovirus co-infection was substantially greater than would be expected if there were no interaction between the viruses. When we did not allow co-transmission from co-infected mosquitoes in our model, the cumulative incidence of co-infection was 22/1,000 individuals per year, a substantial reduction from 46/1,000 individuals per year, the cumulative incidence when co-transmission is allowed. Further, we found that the prevalence of co-infection in our model was always greater than the product of the prevalences of the two individual viruses (Fig. 7). This suggests that these viruses have a synergistic relationship brought on by the presence of co-transmission in the model.

**Figure 7:**
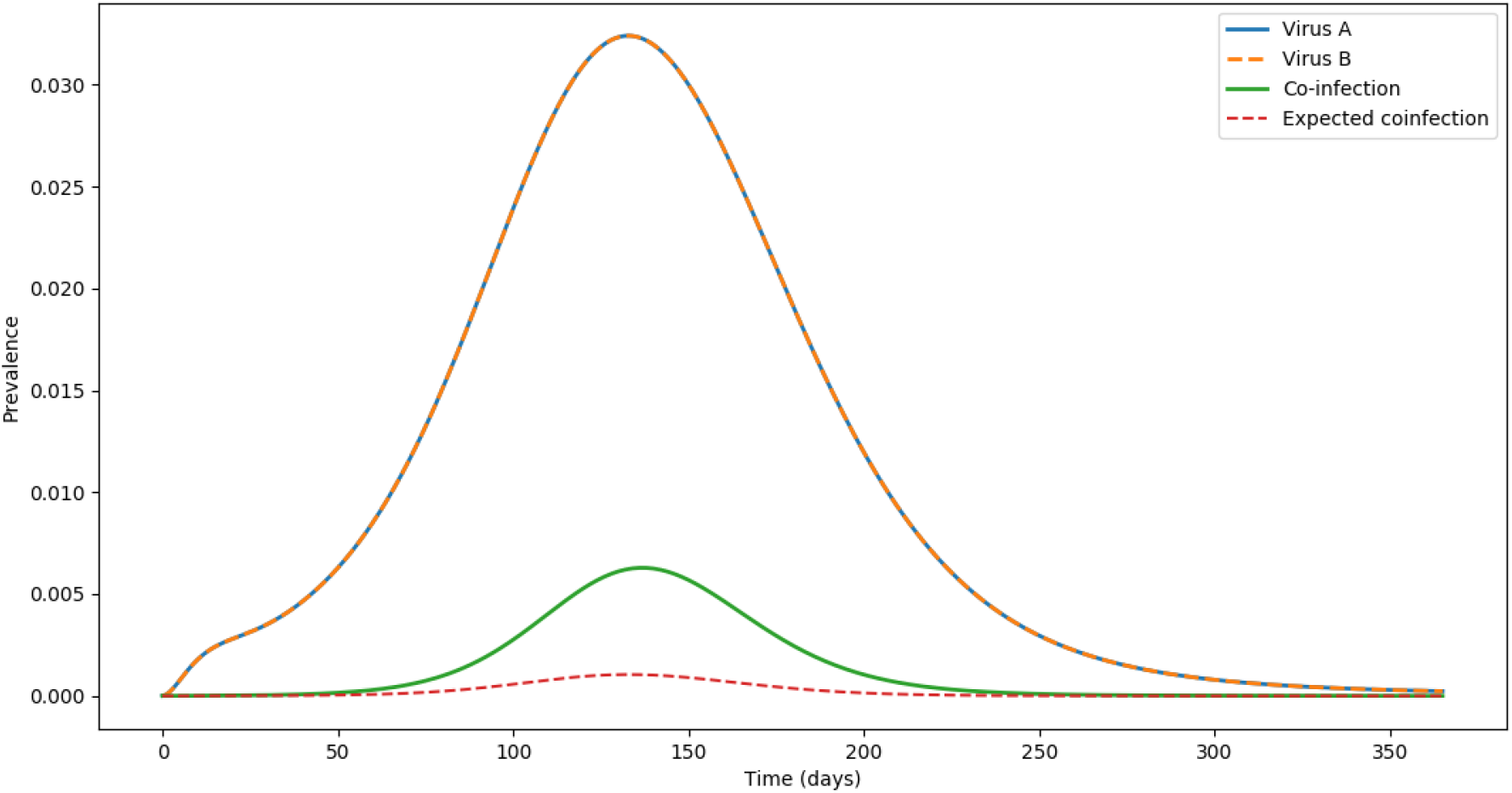
Prevalence of virus A, virus B, observed co-infections, and expected co-infections throughout a year-long simulation of the nonseasonal model, using baseline parameter values. Expected co-infections are defined as the product of the individual prevalences of virus A and B at each time point.

## DISCUSSION

By incorporating seasonal temperature variation, differential importation times, and virus co-transmission, the model implemented here explores the drivers of arbovirus co-infection and assesses conditions under which increased arbovirus co-infection may be likely. To observe substantial co-infection incidence, our model suggests a need for both the consistently favorable temperatures typical of the tropics as well as temporal synchrony between the viruses. In more temperate regions co-infections of the studied arboviruses are rare, only occurring during summer months and even then at very low levels. Repeated seasonal arbovirus outbreaks could result in some degree of cross-protective natural immunity in populations living in tropical environments (13,36,37), which adds an additional layer of complexity to the processes modeled here. Our results suggest that such preexisting immunity to one or more arboviruses could inhibit significant co-infection incidence even when environmental and temporal circumstances are otherwise ideal. However, regardless of immunity, sensitivity analyses indicate that parameters related to seasonality and timing were the primary contributors to variance in model outputs.

This study provides a novel exploration of temperature variation in mosquito life traits and co-infection dynamics within a single modeling framework. Previous modeling studies have characterized the relationship between temperature and arbovirus transmission and have emphasized that warm climates and highly variable moderate climates have high epidemic suitability (28,38). Our results concur with this, and further show that, in some cases, seasonal temperature variation can drive changes in co-infection incidence far more than temperature-independent population parameters do (Fig. 3). As climate change and human mobility patterns lead to expanded arbovirus vector ranges (6), arbovirus co-infection may affect increasing proportions of the global population. The association between higher *Aedes-*borne disease incidence and higher poverty levels (7) indicates that this could become the subject of humanitarian concern, to the extent that co-infections might be associated with more severe outcomes. Increased clinical testing for multiple arboviruses, even if one positive diagnosis has already been obtained, is necessary to provide more informative data on this spread in the future.

We made several assumptions to support parsimonious and computationally tractable model scenarios. First, while temporal synchrony between arriving arboviruses was a crucial component of this study, it is perhaps more likely that an arbovirus would be introduced to an environment where another arbovirus is already endemic, as has been noted in studies of Zika virus and endemic dengue in the Americas (15,39). We explored the dynamics of these scenarios by imposing preexisting immunity to the viruses in turn, as well as simultaneously, and found that immunity to one or both viruses reduces the incidence of co-infection, particularly in the presence of substantial cross-immunity. A similar outcome might be expected in real-world populations where an arbovirus is endemic, although this reduction in incidence of co-infection could be limited if cross-protection is incomplete. Conversely, situations where two novel viruses invade in quick succession, as was observed with Zika and chikungunya viruses in South America in 2013-14 (40) could increase the incidence of co-infection, as model simulations showed here.

Second, for the sake of simplicity, we assumed that the population was homogenous and well-mixed. This could lead to an overestimation of transmission, as high-risk clusters are not always localized both in time and space (17). Incorporating heterogeneity reduces the herd immunity threshold for a population and can lower the arbovirus reproduction number (R_0_), as well (41,42). Within our model, this could reduce the attack rate and influence the ratio of co-transmitted co-infections to sequentially-transmitted co-infections. Finally, model parsimony also influenced our choice of a single mosquito population scaling factor, γ(T), a parameter used to ensure that mosquito populations remained within a reasonable range. While this neglects many complexities of mosquito population dynamics, our use of simplifying assumptions more generally made it possible to isolate the effects of parameters of interest in a straightforward way. Sensitivity analysis of our model allowed us to further examine all parameter interactions and explore the full parameter space.

In this study, we built upon existing modeling work (8) to explore a range of possible influences on arbovirus co-infection through a largely theoretical lens. Expansions of this analysis in the future could benefit from incorporating the growing body of empirical studies exploring the biological mechanisms and outcomes of arbovirus co-infection, particularly those investigating cross-protection and antibody-dependent enhancement (13–16). Data on the frequency of arbovirus co-infection during overlapping epidemics could be informative to this model as well, as could varying parameters between the modeled viruses. While arbovirus co-infection remains a growing area of study, significant work has been done on the interactions and outcomes of a variety of other co-infections, from HIV and tuberculosis (43) to respiratory viral co-infections (44,45). As such, understanding the dynamics of co-infecting pathogens and the clinical consequences of co-infection, especially in the context of global change, is of growing importance for disease mitigation and human health around the world.

## Data Availability

All model code will be uploaded to GitHub in due course

## ACKNOWLEDGEMENTS

This work was supported by the NIH National Institute of General Medical Sciences R35 MIRA program to TAP (R35GM143029) and a Richard and Peggy Notebaert Premier Fellowship from the University of Notre Dame to MLP.

## SUPPLEMENTARY MATERIALS

**Figure S1:**
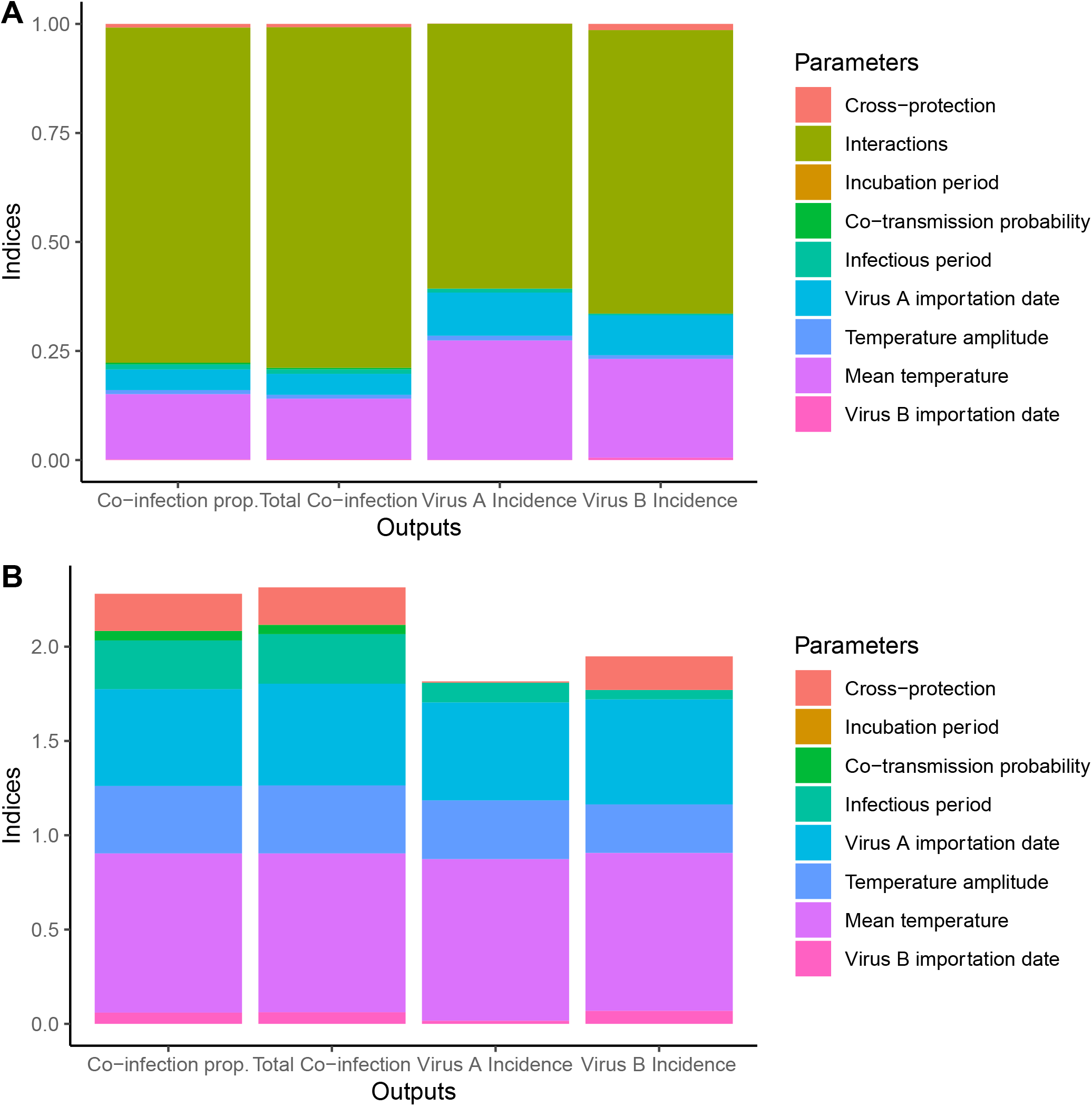
Output of variance-based sensitivity analysis for virus A immunity scenario (50% preexisting immunity to virus A). Both first order (A) and total order (B) indices are shown.

**Figure S2:**
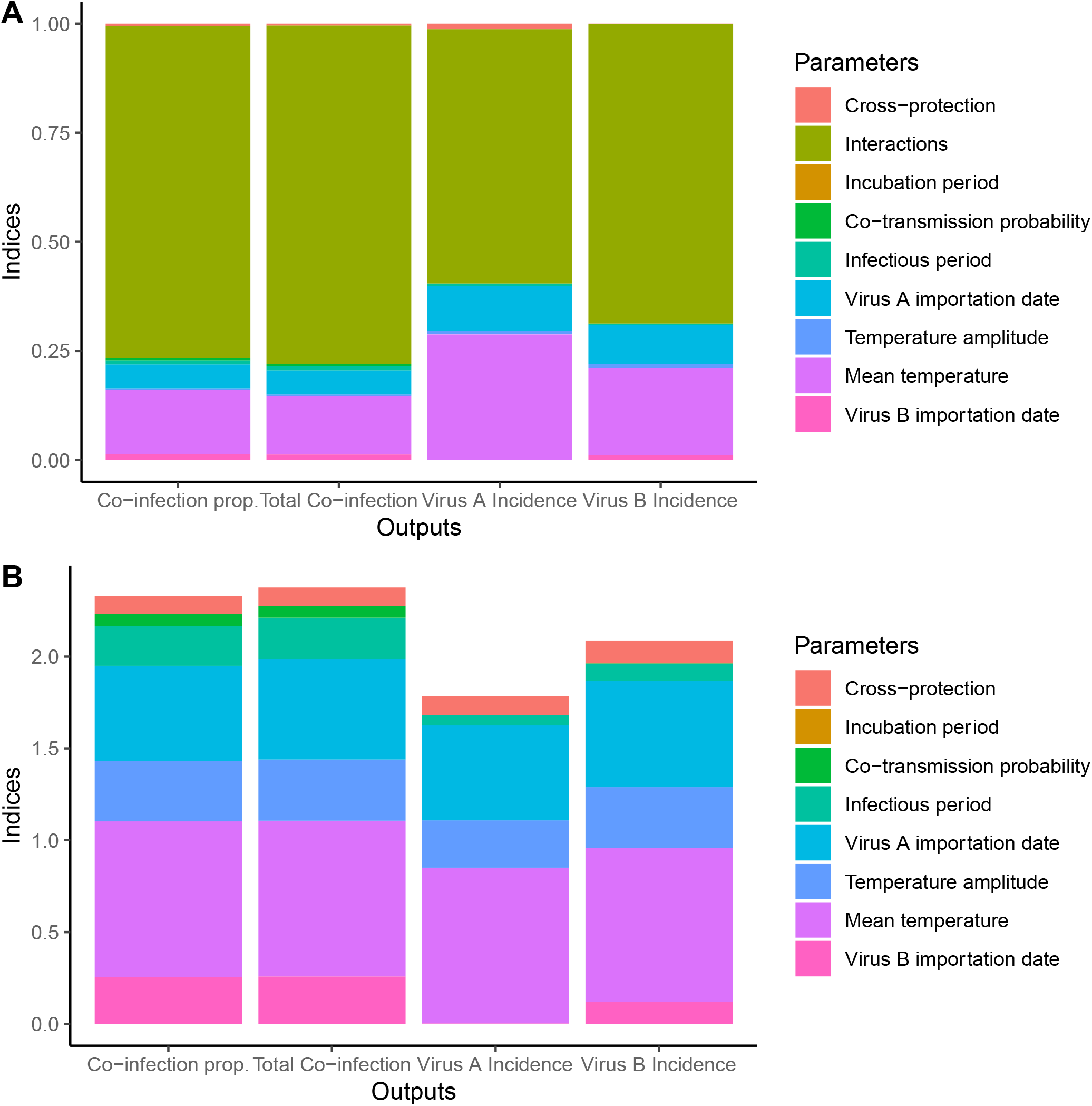
Output of variance-based sensitivity analysis for virus B immunity scenario (50% preexisting immunity to virus B). Both first order (A) and total order (B) indices are shown.

**Figure S3:**
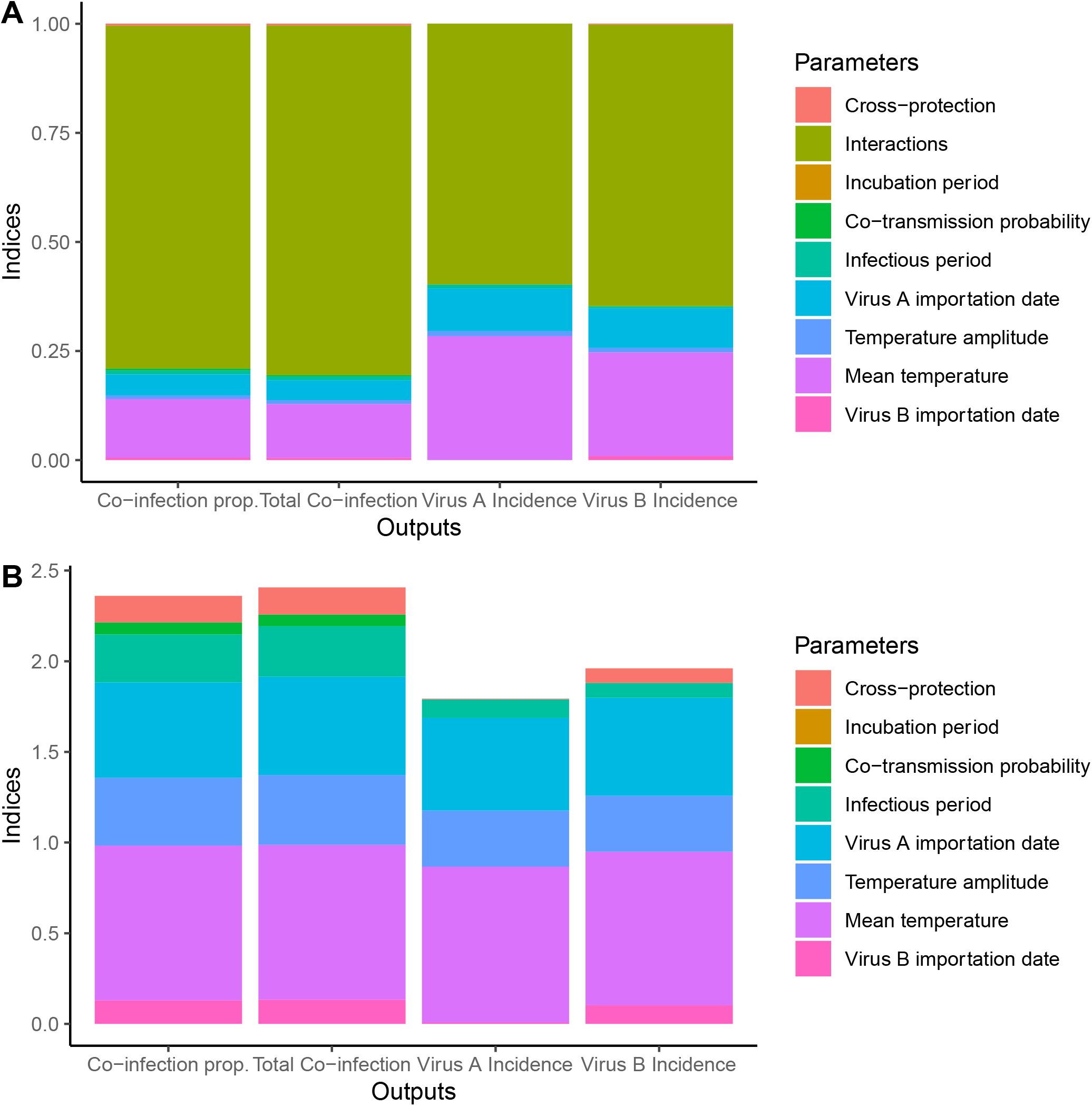
Output of variance-based sensitivity analysis for dual virus immunity scenario (50% preexisting immunity to both viruses). Both first order (A) and total order (B) indices are shown.

